# Global and National Declines in Life Expectancy: An End-of-2021 Assessment

**DOI:** 10.1101/2022.01.14.22269109

**Authors:** Patrick Heuveline

## Abstract

Timely, high-quality mortality data have allowed for assessments of the impact of Covid-19 on life expectancies in upper-middle- and high-income countries. Extant data, though imperfect, suggest that the bulk of the pandemic-induced mortality might have occurred elsewhere. This article reports on changes in life expectancies around the world as far as they can be estimated from the evidence available at the end of 2021.

The global life expectancy appears to have declined by .92 years between 2019 and 2020 and by another .72 years between 2020 and 2021, but the decline seems to have ended during the last quarter of 2021. Uncertainty about its exact size aside, this represents the first decline in global life expectancy since 1950, the first year for which a global estimate is available from the United Nations.

Annual declines in life expectancy (from a 12-month period to the next) appear to have exceeded two years at some point before the end of 2021 in at least 50 countries. Since 1950, annual declines of that magnitude had only been observed in rare occasions, such as Cambodia in the 1970s, Rwanda in the 1990s, and possibly some sub-Saharan African nations at the peak of the HIV/AIDS pandemic.

---

Period life expectancy at birth [life expectancy thereafter] is the most-frequently used indicator of mortality conditions. More broadly, life expectancy is commonly taken as a marker of human progress, for instance in aggregate indices such as the Human Development Index (United Nations Development Programme 2020). The United Nations (UN) regularly updates and makes available life expectancy estimates for every country, various country aggregates and the world for every year since 1950 (Gerland, Raftery, Ševčíková et al. 2014), providing a 70-year benchmark for assessing the direction and magnitude of mortality changes.

Analyses of timely, high-quality vital statistics from about 40 upper-middle- and high-income nations have already demonstrated declines in life expectancies between 2019 and 2020 (Aburto et al. 2021; Islam et al. 2021). Due to the relative efficiency of their mortality reporting, these countries (mostly European, plus the USA and a few countries in East Asia and Oceania) do account for a substantial share of the global deaths attributed to Covid-19 to date. In other countries, however, deaths due to Covid-19 may be more frequently misdiagnosed and under-reported and pandemic-mitigation policies might have induced greater changes in deaths from other causes. Numbers of “excess deaths”—the difference between the actual number of deaths and the number of deaths expected to have occurred in the absence of the pandemic (based on pre-pandemic trends)—would provide a fuller account of the mortality impact of the pandemic (Helleringer and Lanza Queiroz 2021). While imperfect, extant estimates suggests that the number of excess deaths might be two to four times the number of deaths officially attributed to Covid-19 and that the bulk of these excess deaths likely occurred outside of Europe and the other high-income nations in which the mortality impact of the pandemic has been extensively documented (*The Economist* 2022).

This paper presents an attempt to redress this geographical imbalance between the severity of the pandemic and the depth of the current analytical record, by providing estimates of changes in life expectancies up to the end of 2021 for the world and for as many countries as even partial data allow. First, to provide a sense of magnitude for the results, past instances of life expectancy declines are provided from a review of the UN time series from 1950 to 2019. Second, for each country and each quarter of 2020 and 2021, numbers of excess deaths are estimated. Used in combination with previously (pre-pandemic) estimated UN life tables, these numbers yield global and national life expectancies for eight 12-month periods ending each quarter from March 31^st^, 2020, to December 31^st^, 2021. Changes between two consecutive 12-month periods and cumulative 2019-2021 changes are then compared to the UN annual series. The last sections discuss the current data limitations, the estimates’ uncertainty and what might still be reasonably concluded from this still preliminary assessment of global mortality trends between 2019 and 2021.

## Background

### Global life expectancy and mortality crises, 1950-2019

Global and national trends in life expectancies are assessed first to provide context for the pandemic-induced changes. As estimated by the UN, the post-1950 trend in global life expectancy is quite remarkable. The UN estimates that the annual value of the global life expectancy has increased without interruption from 45.7 years in 1950 to 72.6 years in 2019 (United Nations 2019), a .39-year gain per year on average. The largest annual gains, more than .7 year from 1964 to 1968, reflect the success of global public health campaigns, in particular childhood vaccination programs (Cutler, Deaton, and Lleras-Muney 2006).

The distribution of these mortality declines over the lifespan contributed to reduce the global life table entropy (Keyfitz 1977; Goldman and Lord 1986; Olshansky, Carnes, and Désesquelles 2001). As a result, proportionally larger mortality declines would have been required to maintain the pace of annual gains in life expectancy. Instead, annual gains in global life expectancy have gradually declined below their 1950-2019 average of .39 years, dipping below .3 years from 2015 to 2018 and below .2 years in 2019. Annual gains had previously dropped under .2 years between 1990 to 1995, due to HIV/AIDS pandemic, with .16 years in 1992 being the smallest annual gain of the entire 1950-2019 period.

At the national level, countries did not all enjoy an uninterrupted upward trend in life expectancy. Instances of life expectancy declines, from one calendar year to the next, remain rare in the UN time series and relatively modest though. The main exceptions to this generalization are found for Cambodia (up to -4.63 years per year) and Rwanda (up to -5.02 years per year)—two countries that experienced massive increases in violent mortality, in the late 1970s and early 1990s respectively—and a few sub-Saharan countries during by the HIV/AIDS pandemic. According to the UN estimates, the impact of AIDS mortality on life expectancy was most severe in Eswatini in the late 1990s (up to -2.10 years per year).

The UN annual estimates of life expectancy are derived from 5-year period estimates though. This involves a smoothing function that reduces annual variations. The decline in life expectancy in Rwanda between 1993 and 1994, the year of the genocide (Verwimp 2004), is likely much more than five years. National estimates similarly smooth out the impact of mortality crises at the subnational level, such as in Darfur (Hagan and Palloni 2006). Sadly, instances of massive short-term mortality increases driven by violence or famine have not been that uncommon since 1950 (Obermeyer et al. 2008). Contrary to estimates of the number of deaths, however, estimates of life expectancy during these mortality crises remain relatively few. A full reconstruction of demographic changes in China between 1958 and 1961 does suggest that life expectancy may have declined by 12 years between fiscal years 1957-58 and 1958-59 (equivalent to a five-level change in Coale-Demeny model life tables, Ashton et al. 1984: 639). Another reconstruction of demographic changes in Cambodia during the “Khmer-Rouge” regime (1975-78) suggests that life expectancy may have fallen to 8.1 years for males and 16.7 years for females (Heuveline 2015: 211), implying a decline from pre-1975 levels that numbers in decades rather than in years. The conclusion from a review of the UN time series that annual declines in life expectancy since 1950 rarely exceeded two years must thus be qualified as not applying to famine- or violence-driven mortality shocks.

### Pandemic-induced changes in life expectancy

Life expectancy is derived from sex- and age-specific rates of all-cause mortality. The process of verifying and consolidating deaths data to produce these rates is typically a lengthy one. The US Centers for Disease Control and Prevention (CDC), for instance, first produced a provisional estimate of the US life expectancy for 2020 in July 2021 suggesting a decline of 1.5 years compared to 2019 (Arias et al. 2021). The estimated decline was increased to 1.8 years with the final estimate released in December 2021 (Murphy et al. 2021). Given the urgency to document recent mortality conditions during the pandemic, provisional mortality statistics have been released notably faster than under usual circumstances. Most notably, the Human Mortality Database has released a Short-term Mortality Fluctuations data series (STMF, Jdanov et al. 2021) that tracks weekly mortality data with a few-week lag for countries with reliable and timely mortality statistics. Analyses of these data have provided estimates of life expectancy change in 2020 for nearly 40 countries, mostly European, with a few additional upper-middle- and high-income nations in North America, East Asia and Oceania (Aburto et al. 2021; Islam et al. 2021). From these analyses, the only country that appears to have experienced a 2-year or larger decline in life expectancy between 2019 and 2020 is Russia. (Islam et al. 2021 report the difference between the actual and expected 2020 values, 2.33 for males and 2.14 for females, which should be slightly larger than the 2019 to 2020 decline given the expected, counterfactual, upward trend).

For all but a handful of countries, numbers of COVID-19 deaths that are updated at least daily on online dashboards such as Johns Hopkins University’s (JHU) have provided a timelier and, foremost, more global resource to assess the mortality impact of the pandemic (Dong, Du and Gardner 2020). Analyses of these data strongly suggest that the largest declines in life expectancy were not occurring in Europe or the USA, but in countries of Central and South America (Heuveline and Tzen 2021). The life expectancy estimates derived from these data proved unreliable, however, as their validity depends on reported counts of deaths due to Covid-19 that may be inaccurate and an assumption of unchanged rates of mortality from causes other than Covid-19 that may not hold. Cause-specific mortality data reveal increases in US death rates from causes other than Covid-19 during the pandemic for instance (Ahmad and Anderson 2021). Conversely, analyses of STMF data showed that, in a few countries, life expectancy increased more in 2020 than in recent years before, suggesting that public health interventions intended to mitigate the impact on the virus also reduced mortality from other causes. With respect to deaths attributed to Covid-19, protocols that require including all suspected, but unconfirmed, Covid-19 deaths might have produced overcounts in some countries (Beaney et al. 2020). The main concern, however, remains the possibly vast extent to which Covid-19 deaths might have been misdiagnosed or unreported in many parts of the world. Estimates of excess deaths in Central and South America suggest drastically larger reductions in life expectancy than when based on reported Covid-19 deaths, reaching 10.91 years in Peru, 7.91 years in Ecuador, 5.54 years in Mexico, 2.42 years in Brazil, and 2.26 years in Guatemala (Lima et al. 2021).

## Data and Methods

### Excess deaths data

The most comprehensive source of excess-death estimates to date is the *World Mortality Dataset* (WMD), which at the end of 2021 covered over 100 countries (Karlinsky and Kobak 2021). In the combined population of these countries, the WMD suggests there were 60% more excess than Covid-19 deaths since the beginning of the pandemic. However, the scope and quality of the mortality data available to estimate excess mortality varies across countries. An analysis by *Our World In Data* (OWID) analysts suggests that nearly 60 of these countries, the data did not allow for reliable estimation of the expected number of deaths over time from which estimates of excess deaths are derived (OWID 2021). Reasons include too few years of pre-pandemic data to estimate the temporal trend, insufficient breakdown over time (within year) to adjust for seasonality, or insufficient age breakdown to adjust for demographic changes.

Even the full WMD still does not cover large swaths of Africa and in Asia. The few sub-Saharan African countries that are included (Mauritius, Mayotte, Reunion, Seychelles and South Africa), for instance, are clearly not representative of the entire region. But the most conspicuous coverage gap and current unknown quantity may be India. A recent study derived from three independent data sources estimated a confidence interval of 2.75 to 12.25 for the ratio of excess to Covid-19 deaths (Anand, Sandefur, and Subramanian 2021). Culling mortality data from several sources, however, the most sophisticated demographic analysis to date suggested the number of excess deaths was likely seven times the official number of Covid-19 deaths at the time (Guilmoto 2021). This ratio is consistent with the largest study of the Civil Registration System that placed the number of excess deaths close to 3 million by the end of 2021, more than six times the official tally at the time (Jha et al. 2022).

A machine learning algorithm designed to provide estimates of excess deaths for all countries and the whole world suggests that the global number of excess deaths from the start of the pandemic to the end of the 2021 is between 2.2 and 4 times the reported number of Covid-19 deaths (*The Economist* 2022). The algorithm (known as “gradient boosting”) is developed by fitting the relationship between excess mortality and a large set of diverse national indicators (including mean elevation, average temperature, and prevalence of malaria, TB or HIV) on a training sample of about 80 countries. This model provides estimates of excess mortality for many countries where there is no reliable Covid-19 mortality data for such estimation—or only unrepresentative data, typically from small studies conducted in urban centers (e.g., Jakarta, Indonesia: Djaafara et al. 2021; Khartoum, Sudan: Watson et al. 2020; Damascus, Syria: Watson et al. 2021; Lusaka, Zambia: Mwananyanda et al., 2021; Aden, Yemen: Koum Besson et al. 2021). As with any extrapolation strategy, however, the performance of gradient boosting algorithms depends on the degree of similarity between countries included in the training sample and other countries.

### An estimate of global excess deaths

Numbers of excess deaths were estimated for each country and each quarter in 2020 and 2021. Different approaches were used depending on the availability and quality of the data in each country. For a first group of 53 WMD countries, data quality was deemed satisfactory per OWID criteria. In these countries, WMD estimates of excess deaths by quarter were used when available for the entire quarter. In the remaining quarters, excess deaths were estimated iteratively based on the relationship between estimates of excess deaths and Covid-19 deaths reported on the JHU dashboard in the past 12 months. When these excess-death estimates were larger than reported Covid-19 deaths, the ratio of the two was applied to the number of Covid-19 deaths reported on the JHU dashboard for the following quarter. This assumption would fit the situation where excess deaths are mostly due to Covid-19 and the ratio of reported to unreported Covid-19 deaths does not change. When excess-death estimates over the past 12 months were less than reported Covid-19 deaths, or even negative, the average quarterly difference between the two was assumed to remain the same in the next quarter. This assumption would fit the situation where Covid-19 deaths are accurately reported and deaths from other causes are fewer than expected by the same amount each quarter.

The second group of countries includes the remaining WMD countries plus some, like India, for which a national estimate of excess mortality might be available from ancillary sources. For the WMD countries, preliminary quarterly estimates were produced following the same approach as for the first group of countries. The end-of-2021 tallies of excess deaths were then compared to the number derived from the tallies of Covid-19 deaths reported on the JHU dashboard and the percent undercount predicted by *The Economist* model. In countries for which preliminary estimates already exceeded reported Covid-19 deaths in each quarter, and the predicted undercount suggested an even higher ratio of excess to Covid-19 deaths, the preliminary quarterly estimates were scaled up to match the predicted undercount. In other countries, the undercount ratio predicted by the model or provided by ancillary sources was applied to quarterly numbers of Covid-19 deaths reported on the JHU dashboard.

This last approach is the only option to estimate excess deaths in the remaining countries comprising nearly half of the world population. Since the performance of the model remains difficult to assess at this point for countries for which there has been little to no direct data on Covid-19 mortality, an upper limit was placed on the predicted cumulative number of excess deaths. This limit was set by deriving age-standardized rates of excess mortality for the 2020-2021 period for each country within each UN geographic areas, and if necessary, scaling down the prediction for a country in this third group so that its rate would not exceed the highest age-standardized rate for countries in the first two groups. The full calculations for each country are provided in Supplementary File (“Excess Deaths”).

Across groups, this estimation yields more than 15 million excess deaths in 2020 and 2021, 2.8 times the global number of Covid-19 deaths reported at the end of 2021 (5.4 million). Figure 1 summarizes the estimated numbers of excess deaths for each group of countries and the global number of reported Covid-19 deaths for each quarter in 2020 and 2021. The Figure shows that the exact proportion by which reported Covid-19 deaths underestimate excess deaths worldwide depends largely on the situation in countries where it has only been partially (Group II) or hardly (Group III) documented. As also illustrated in Figure 1, excess mortality trends differ across Groups. While 33% of excess deaths occurred in Group-I countries in 2020, only 24% of 2021 excess deaths occurred in these countries. Global trends cannot be simply extrapolated from the well-documented trends in Group-I countries.

**Figure 1:**
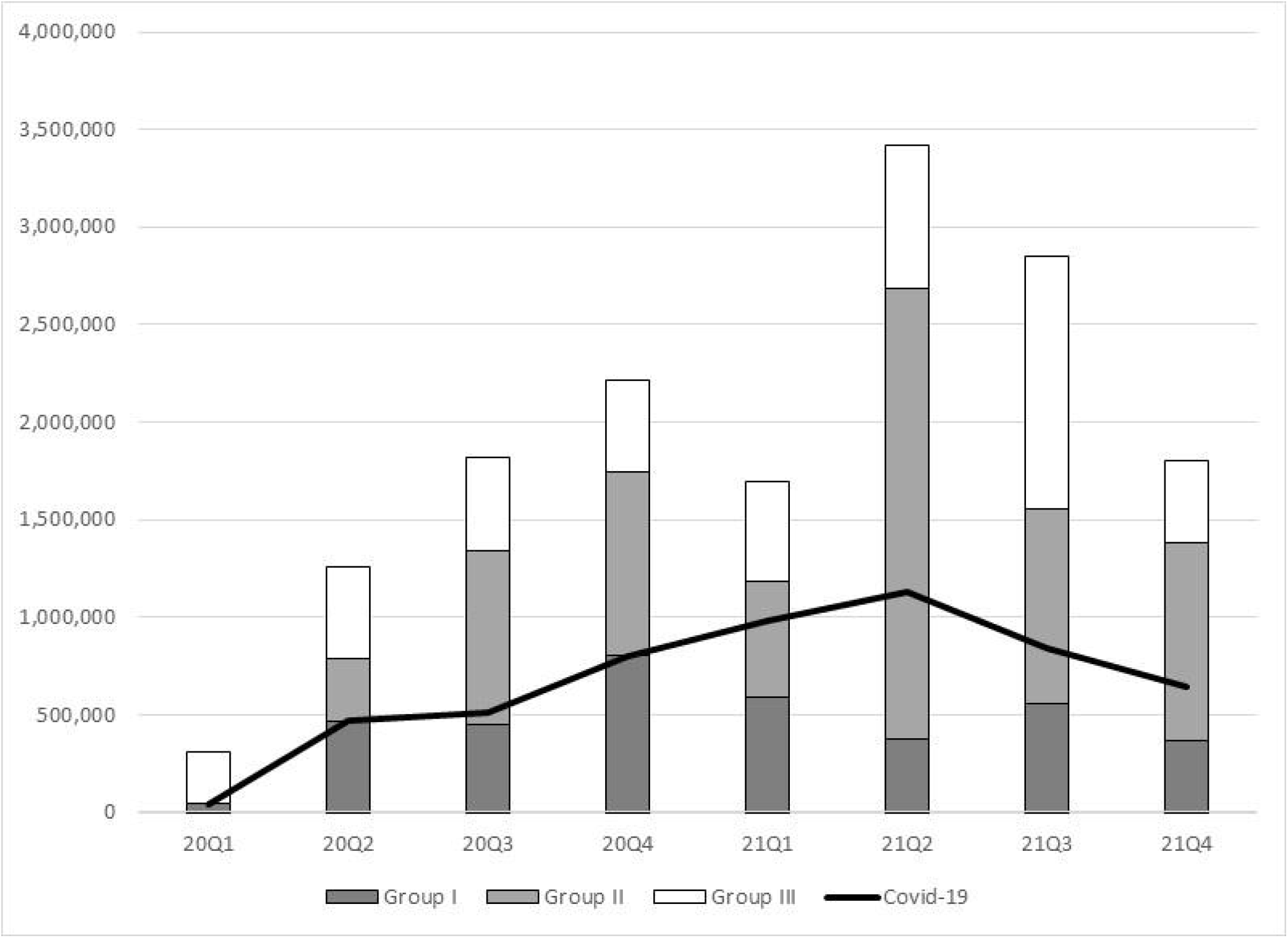
Estimates of excess deaths by country group and reported Covid-19 deaths, 2020-21, by quarter. Note: see text for definition of Group-I, Group-II & Group-III countries. Sources: Covid-19 deaths, JHU online dashboard; Excess deaths, author’s calculations (see supplementary files for details)

### Methods: Recalculating period life expectancies

Life expectancies were recalculated for eight 12-month periods, each ending in one of the quarters of 2020 and 2021 (the first period being from April 1^st^, 2019, to April 1^st^, 2020, and the last one being the calendar year 2021). The estimation of global and national life expectancies in each period proceeded in four steps. First, excess deaths were distributed by age and sex. A different sex- and age-pattern was used in each period, based on the cumulative number of COVID-19 deaths by sex and age-group reported by the US Centers for Disease Control and Prevention (CDC) at each quarter end (CDC 2022). The number of excess deaths in each sex- and age-group was derived from the total number of excess deaths in the world/country during a period, the number of COVID-19 deaths in the same sex-age-group and period in the USA and the ratio of sex- and age-group’s population size in the world/country and in the USA. That last ratio was obtained from the UN projections of national population sizes by sex and age-group for mid-2019, -2020 and -2021 (United Nations 2019).

Second, sex- and age-specific mortality rates (_*n*_*m*_*x*_) and survival probabilities (_*n*_*p*_*x*_) for the calendar year 2019 and counterfactual rates and probabilities for each of the eight 12-month periods were derived from the UN 2019 estimates and projections. Period values were obtained by linear (_*n*_*m*_*x*_) or exponential (_*n*_*p*_*x*_) interpolation between the 2015-20 and 2020-25 values from the UN. Third, excess mortality rates for each country and the world in each of the six periods were combined with the counterfactual rates into period life tables by reversing the procedure typically used to “delete” a cause of death from a multiple-decrement life table (Preston, Heuveline, and Guillot 2001; Heuveline and Tzen 2021). Fourth, life expectancies were estimated from the re-estimated probabilities and the counterfactual rates and probabilities. Additional details on these four steps are provided in the Appendix (steps 2 to 5). National life expectancies were only estimated for all Group-I and Group-II countries for which the UN estimates life table functions (countries with a population size above a given threshold)—a total of 98 countries. Estimates of excess deaths for Group-III countries were not deemed sufficiently reliable for life expectancy estimation.

## Results

The increase in the number of deaths during the pandemic had a substantial impact on the global life expectancy. After 69 years of uninterrupted increase from 1950 to 2019, the global life expectancy is estimated here to have declined by -.92 years between 2019 and 2020 (for both sexes), and by another .72 years between 2020 and 2021 (Figure 2). In 2021, global life expectancy is estimated to have dropped below its 2013 level.

**Figure 2:**
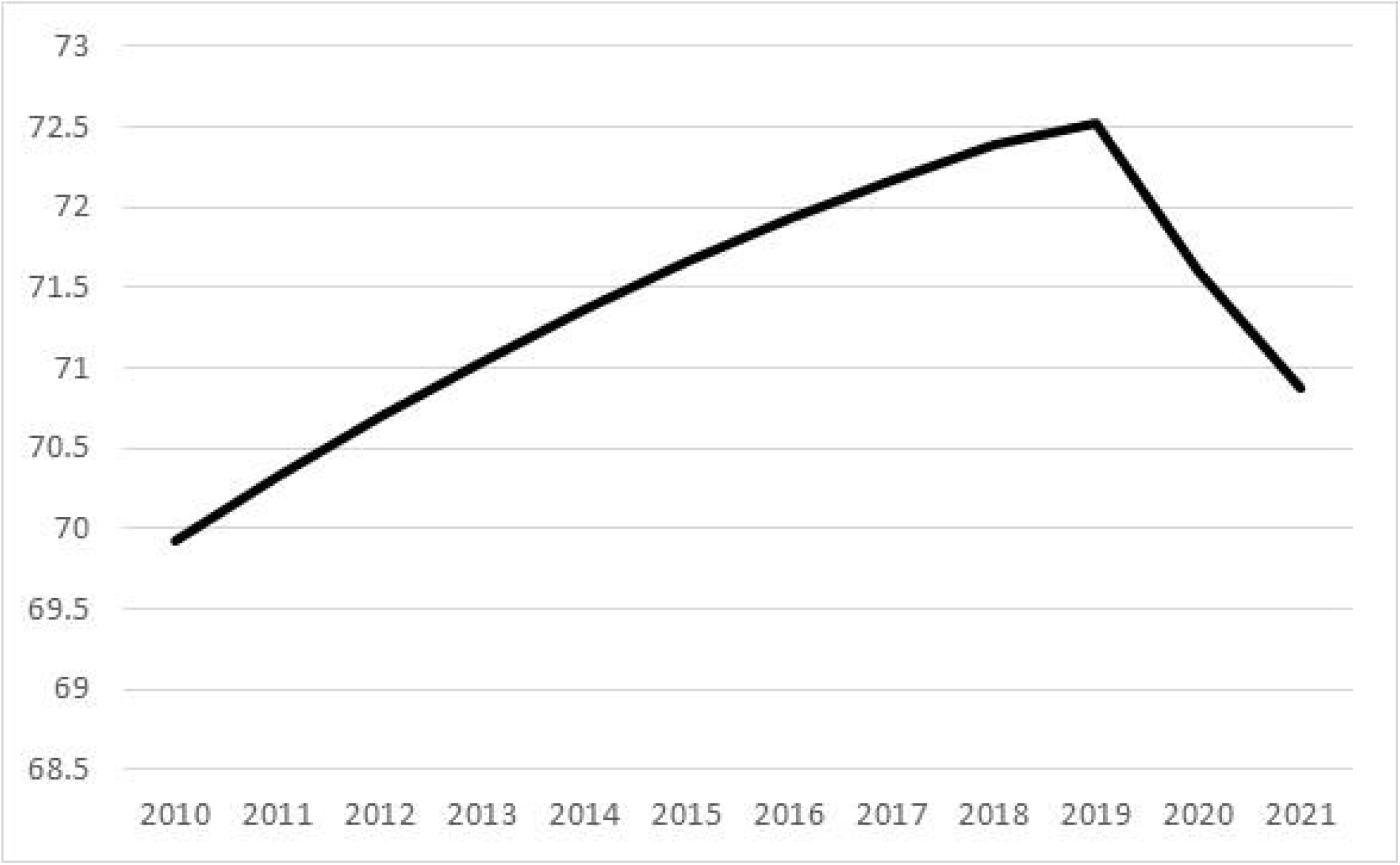
Global life expectancy, 2010-21 (both sexes, in years) Sources: 2010-19, United Nations (2019); 2010-21, author’s calculations (see Appendix for details)

Comparing life expectancy estimates for each of the eight 12-month periods, however, the decline in global life expectancy appears to have stopped in the last quarter of 2021 (Figure 3). Based on these eight estimates, tracking changes in life expectancy between two consecutive 12-month periods (annual change thereafter) shows that the annual change for the global population is estimated to have peaked at 1.33 years at the end of June 2021 (mid-2020 to mid-2021 v. mid-2019 to mid-2020).

**Figure 3:**
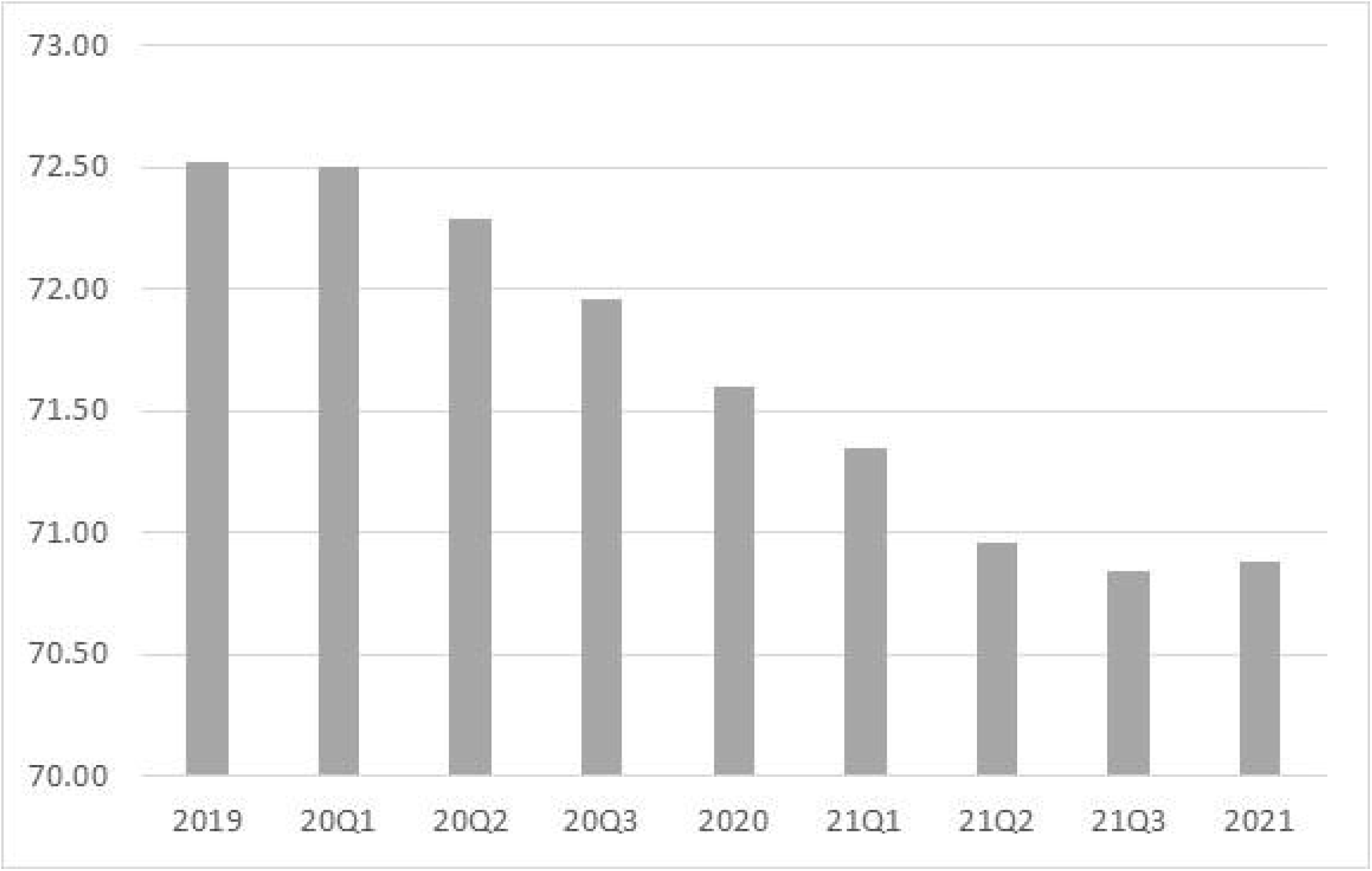
Global life expectancy, by 12-month period ending in each quarter of 2020 & 2021 (both sexes, in years) Note: *YQn* refers to the 12-month period ending at the end of the n^th^ quarter of year 2000+*Y* (e.g., 20Q1 is the period including the last three quarters of 2019 and the first quarter of 2020). Sources: 2019, United Nations (2019); 2010-21, author’s calculations (see Appendix for details)

At the national level, many countries experienced substantial changes in life expectancy (Figure 4). Between 2019 and 2021, life expectancy is estimated to have declined by more than two years annually (four years overall) in eight countries (Figure 4, category 3), five in America (Peru, 5.6, Guatemala, 4.8, Paraguay, 4.7, Bolivia, 4.1, and Mexico, 4.0 years) and three in Europe (the Russian Federation, 4.3, Bulgaria, 4.1, and North Macedonia, 4.1 years).

**Figure 4:**
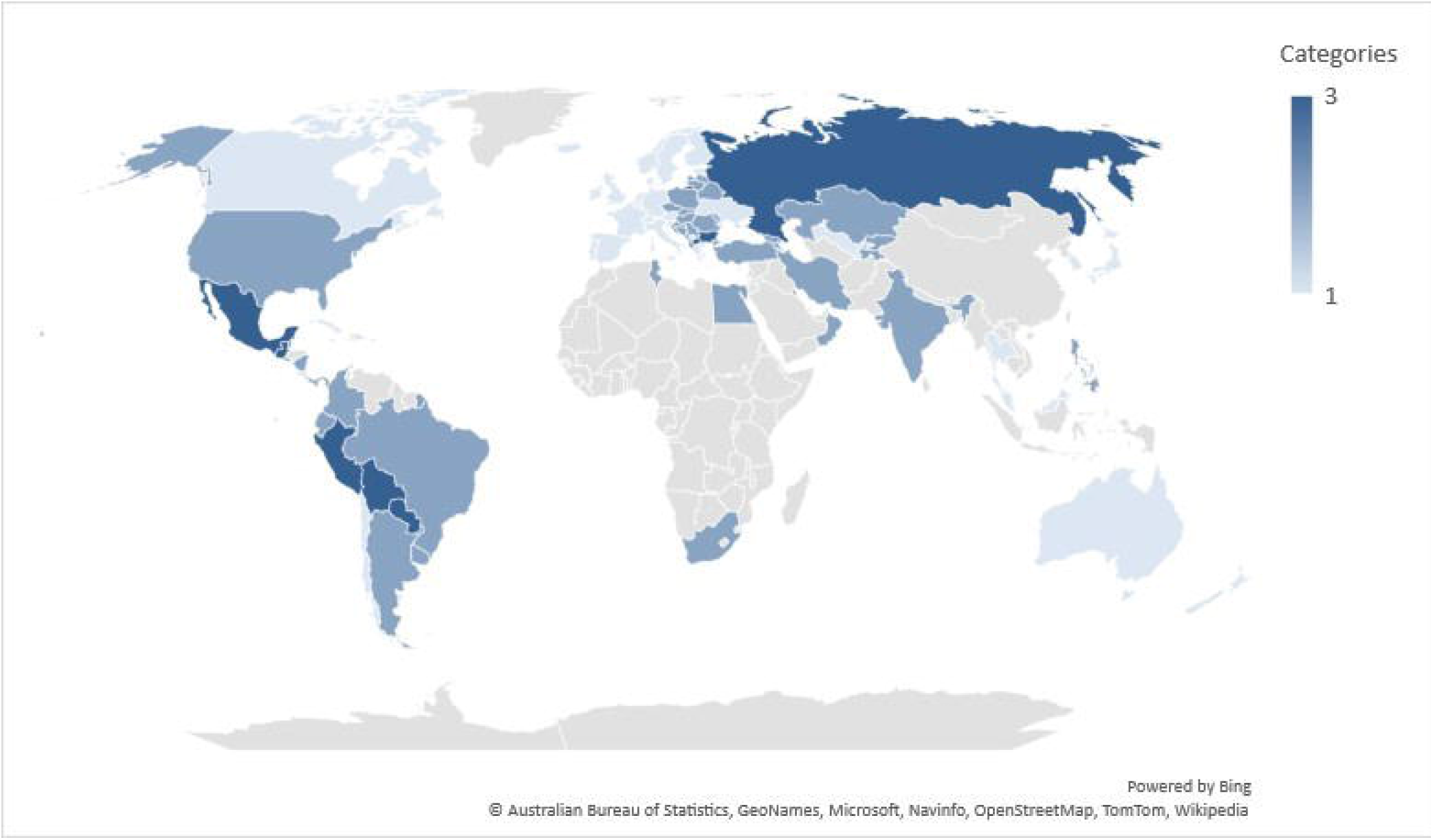
Annual change in life expectancy, 2019-21 (both sexes, in year) Categories: 1: Maximum annual decline < 2 years 2: Maximum annual decline > 2 years, average annual decline <2 years 3: Average annual decline >2 years Sources: Author’s calculations (see supplementary files for details)

Tracking annual change at the end of each quarter, however, more than half of the countries for which life expectancies were estimated (53 out of 98) reached an annual change in excess of two years at some point in 2020 or 2021 (Figure 4, category 2). Annual change even reached seven years in Peru and between four and six years in several other countries in America (Mexico, Nicaragua, Bolivia, Paraguay, Columbia, Ecuador and French Guiana). In Europe, annual change reached a little over four years in Bosnia and Herzegovina and in North Macedonia and over three years in few other countries (Montenegro, Bulgaria, Albania, and Poland). Substantial annual changes are also observed throughout Asia, from Southeast Asia (Philippines, 3.0 years) and South Asia (India, 2.6 years) to Central Asia (Kazakhstan, 3.2 years) and Western Asia (Lebanon, 3.4 years), and in the few countries in continental Africa with sufficient data (Tunisia, 3.4 years, South Africa, 3.1 years, and Egypt 2.3 years). Among those with sufficient data, the only countries that did not reach that the two-year mark at any point between 2020 and 2021 are countries in Eastern Asia, Australia, New Zealand and European countries west of a line running from the Baltic to the Balkans. Together with the USA, which did reach an annual change of just over two years, these are arguably the countries where the impact of the pandemic has been the most extensively studied to date.

Tracking change quarterly also reveals very diverse timing of pandemic impact across countries (see Supplementary File “Life Expectancies”). In some (Nicaragua, Ecuador), annual change peaked in 2020 and life expectancy recovered in 2021. On the contrary, after little change in 2020, the annual change was still increasing in the last quarter of 2021 in the Philippines and Overseas Territories of France (Guyana, Martinique and Guadeloupe), for instance. The plateau in global life expectancy reached during the last quarter of 2021 (Figure 3) is far from a global trend and results instead from a diminishing impact of the pandemic in some countries and a still increasing impact on some other countries.

## Discussion

The results demonstrate that the pandemic had an impact on the global life expectancy that has no precedent since 1950. In more than half of the countries where impacts on national life expectancy could be estimated, they also appear to be of a rare magnitude since 1950. Obviously, there is still substantial uncertainty about the exact size of the declines in life expectancy even in these countries and globally. Estimates of declines in life expectancy were derived here from numbers of excess deaths that in turn must be derived from statistical modelling of what the number of deaths might have been in the absence of the pandemic. Even in countries with the required good-quality data, this modelling involves multiple decisions for which there is no clear rule—regarding the number of past years used to define benchmark mortality conditions, if and how a temporal mortality trend is modelled, etc.—and which may substantially impact the results (Nepomuceno et al. 2021). The main challenge to measuring excess deaths with confidence, however, remains substantial data limitations in many parts of the world.

An additional difficulty for some countries is that only a total number of excess deaths might be derived from the number of deaths attributed to Covid-19 and ancillary data, but the impact of these excess deaths on life expectancy depends on their age and sex distribution. As age-and-sex distributions of excess deaths are only available in a limited number of countries, the results presented here rest on a simplifying assumption that derived the distribution of excess deaths in all countries to from a single mortality schedule (US sex- and age-specific mortality rates from Covid-19), albeit different for each period. For countries that have good quality data on excess deaths by age and sex, extant results based on these data should be more reliable than those presented here, the former providing useful benchmarks for assessing the quality of the latter.

In settings where excess deaths consist mostly of reported and unreported Covid-19 deaths, the main issue is expected to be potential differences in sex-and-age patterns of Covid-19 mortality that between countries. Extant reviews suggest that the age patterns are flatter in lower-income countries and at lower life expectancy levels (Demombynes et al. 2021; Guilmoto 2020; Ioannidis,⍰Axfors,⍰and Contopoulos-Ioannidis⍰2021; O’Driscoll et al. 2021). Further evaluations of data quality might be needed to validate that observation, as a higher degree of uncertainty about exact age can also reduce the slope of the mortality schedule (Preston et al 1996). Moreover, even if these observed differences in slope were to be taken at face value, Covid-19 mortality schedules would still be broadly similar (Ohnishi, Namekawa and Fukui 2020). A sensitivity analysis substituting the US sex-and-age pattern of Covid-19 mortality to the pattern prevailing in Brazil, for instance, did produce an “older” distribution than the actual distribution of excess deaths, but only reduced the estimated impact on 2020 life expectancy by 3% (Heuveline and Tzen 2021). Finally, higher vaccination rates at older ages have also resulted in flatter age patterns in high-income nations over time and should have reduced the difference in the concentration of Covid-19 mortality at older ages between countries.

A different issue might be expected in settings where the number of excess deaths is substantially affected by changes in the number of deaths from other causes. Because the distribution of Covid-19 deaths is older than the distribution of deaths from most other causes, one excess death due to Covid-19 has less impact on life expectancy that one excess death from another cause. One study estimated that in 2020 Covid-19 deaths accounted for 83% of excess deaths but only 73% of the years of life lost in the USA for instance (Chan, Cheng and Martin 2021). The 2019-to-2020 decline in US life expectancy estimated here (1.63 years, see Supplementary File “Life Expectancies”) is indeed smaller than the CDC’s final estimate (1.8 years, Murphy et al. 2021). Conversely, in countries where a ratio of excess to Covid-19 ratio below one could reflect lower than expected mortality from causes other than Covid-19 during the pandemic, the expectation is that the impact of excess deaths would be over-estimated. The 2019-to-2020 decline in life expectancy estimated here for France (.70 years, see Supplementary File “Life Expectancies”) is indeed larger than the country’s official estimate (.5 years for females and .6 years for males, Papon and Beaumel 2021).

The sensitivity of the results to differences in age and sex patterns of excess mortality appears relatively modest and in most countries uncertainty about the total number of excess deaths is by far the main concern. Neither the uncertainty about the scale of excess mortality, nor the uncertainty about its distribution by age and sex appear to be substantial enough though to invalidate the finding that the global life expectancy declined in 2020 for the first time in 70 years and continued to decline between 2020 and 2021. With 30% of the global excess deaths being estimated in Group-III countries, the pandemic impact on global life expectancy could be substantially smaller than estimated here, but not to the point that life expectancy would have continued to increase. The figure of 15.4 million excess deaths at the end of 2021, on which global life expectancy estimation rests, is 2.8 times the number of global deaths officially attributed to Covid-19 at that point, when *The Economist* (2022) model provided 2.2 to 4.0 as a 95% confidence interval for that ratio. At the national level, the result that many countries have experienced a decline in life expectancy since the beginning of the pandemic should also be a robust finding. The exact number is difficult to assess, due less to the uncertainty of the estimates presented here than to the fact that, as of this writing, post-pandemic life expectancy could not yet be estimated in roughly half of the countries.

When it can be estimated, interpreting these reductions in life expectancy is not entirely straightforward either. The popularity of life expectancy as a summary indicator of mortality conditions results in part from its intuitive interpretation as an average length of life were mortality conditions to remain unchanged. The meaning of a change in life expectancy driven by hopefully temporary changes in mortality is less intuitive (Goldstein and Lee 2020; Modig, Rau and Ahlbom 2020; Heuveline 2021a). (See Appendix for further discussion of a possible interpretation of temporary changes in life expectancy). Several alternative measures have been proposed to express how much changing mortality conditions have impacted longevity during the pandemic (Ellege 2020; Goldstein and Lee 2020; Verdery, Smith-Greenaway, Margolis, and Daw 2020; Heuveline 2021a; Pifarré i Arolas, Acosta, López Casasnovas et al. 2021). But life expectancy remains the most available summary indicator of mortality conditions across the world and over time, providing unique opportunities for geographic and historical comparisons. In this respect, post-pandemic trends in many countries unambiguously signal a mortality impact at a scale rarely observed since 1950 except during famines and violent conflicts.

Conducting these analyses at the national level is largely a data-driven choice. Several analyses have demonstrated important within-country differences both across geographical units (e.g., Castro et al. 2021; Garciá-Guerrero and Beltrán-Sánchez 2021; Heuveline and Tzen 2021) and between racial/ethnic groups (Andrasfay and Goldman 2021). World maps such as Figure 4 would conceal high impacts on sub-populations in relatively better-off countries.

## Conclusion

Changes in life expectancy between 2019 and 2020 in America, Europe, and a few other countries have received copious attention. Results presented here confirm several key take-aways from previous analyses such as the large mortality impact of the pandemic (1) in the USA relative to other high-income nations in Western Europe (Aburto et al. 2021; Heuveline 2021b), (2) in Russia relative to the rest of Europe (Islam et al. 2021), and foremost, (3) in some Central and South American nations (Lima et al. 2021).

Using end-of-2021 reports of deaths attributed to Covid-19 and modelling their relationship to excess deaths, preliminary estimates were also prepared for changes in life expectancy in 2021. These results suggest a growing gap between, on the one hand, Western European nations and, on the other hand, the USA, where life expectancy continued to decline, and even more so, Russia, where it is expected to decline more than in 2020. In Central and South America, the record is more contrasted with countries where life expectancy is expected to recover some of the large declines of 2020 (e.g., Ecuador, Nicaragua), or to continue to decline but substantially less than in 2020 (e.g. Bolivia, Mexico, Peru) and some where the 2021 declines are expected to exceed the 2020 declines (e.g., Brazil, Columbia, Guatemala, Paraguay).

Changes in life expectancies were also estimated for a total of 98 countries including some that had not received as much attention to date. These results highlight a geographical imbalance between the availability and quality of data on excess mortality and impact of the pandemic. At an early stage in the pandemic, the quantity of data might have been commensurate with the severity of the pandemic. The first wave of the pandemic was well documented as it affected high-income countries with good statistical systems, foremost in Western Europe and the USA (Kontis et al. 2020; Vestergaard et al. 2020). As these analyses have shown, this is no longer the case. With the notable exception of the USA, the annual change in life expectancy in these wealthy forerunners has never reached the level observed in over half of the countries with the data required for sufficiently reliable estimation. The mortality impact of the pandemic has shifted from West to East in Europe, and globally from North to South. As far as current empirical limitations allow them to be quantified, more than 20% of global excess deaths to date might have occurred in India, where an understanding of the scale of the pandemic is slowly emerging, and possibly another 30% in countries where there is hardly any reliable source to evaluate the local situation. As the results suggest substantial mortality reversals in many parts of Asia, and possibly Africa as well, of a magnitude rarely observed since 1950, the need for better monitoring mortality trends in these countries cannot be overemphasized (Helleringer and Lanza Queiroz 2021).

An attempt to estimate the global life expectancy since the beginning of the pandemic despite these data limitations indicated in a .92-year decline between 2019 and 2020 and a .72 decline between 2020 and 2021. By contrast, the UN (2019) anticipated a .18-year gain in global life expectancy between 2019 to 2020. The 2021 global life expectancy would then be two full years before its previously expected level and below its estimated 2013 level. While it is still too early to confidently quantify this decline in global life expectancy, a decline is already beyond doubt, signaling a unique feature of the mortality changes induced by the pandemic. Each year since 1950, years of life lost to mortality reversals in some parts of the world had been more than compensated by years of life gained from declines in other causes of deaths or in other parts of the world. For the first time in at least 70 years, this was not the case in 2020 and will not be the case in 2021 either.

The decline did appear to stabilize at the end of 2021. As has been the case throughout the pandemic, however, the mortality impact was still increasing in some populations while decreasing in others. The seemingly positive trend merely resulted from the fact that at the end of 2021 the increasing impact was mostly observed in comparatively small populations, in archipelagoes in particular. The end-of-2011 trends looked more encouraging that they had in nearly two years, but It would certainly appear unwise at this point to claim that the impact of the pandemic on the global life expectancy has peaked.

## Supporting information

Estimation of Excess Deaths

Life Expectancy Estimates

## Data Availability

All data are from publicly available sources 1) Demographic data (2019, 2020 & 2021 world population by age and sex and counterfactual life tables by sex) from the United Nations: https://population.un.org/wpp/Download/Standard/Interpolated/ 2) Global and national numbers of COVID-19 deaths in 2020 from Johns Hopkins University: https://coronavirus.jhu.edu 3) US number of COVID-19 deaths by age and sex from the Centers for Disease Control and Prevention (CDC): https://data.cdc.gov/NCHS/Provisional-COVID-19-Death-Counts-by-Sex-Age-and-S/9bhg-hcku 4) Excess mortality estimates are from the World Mortality Dataset (WMD) at https://github.com/akarlinsky/world_mortality 5) Covid-19 death undercounts are from The Economist at https://www.economist.com/graphic-detail/coronavirus-excess-deaths-estimates

https://coronavirus.jhu.edu

https://data.cdc.gov/NCHS/Provisional-COVID-19-Death-Counts-by-Sex-Age-and-S/9bhg-hcku

https://github.com/akarlinsky/world_mortality

https://www.economist.com/graphic-detail/coronavirus-excess-deaths-estimates

## Appendix

### Estimating changes in life expectancy

The estimation of global and national life expectancies proceeded in the following steps.

#### Step 1

Estimates of excess deaths as of March 31^st^, 2020, June 30^th^, 2020, September 30^th^, 2020, December 31^st^, 2020, March 31^st^, 2021, June 30^th^, 2021, September 30^th^, 2021, and December 31^st^, 2022 were used to calculate excess deaths in eight twelve-month periods, each ending at one of these dates.

#### Step 2

Excess deaths were distributed by age and sex. A single sex- and age-pattern was used in each period, based on the cumulative number of COVID-19 deaths by sex and age-group reported by the US Centers for Disease Control and Prevention (CDC) for that period (CDC 2022). The number of excess deaths in each sex- and age-group was derived from the total number of excess deaths in the world/country during a period, the number of COVID-19 deaths in the same sex- and age-group in the U.S.A. in that period and the ratio of sex- and age-group’s population size in the world/country and in the U.S.A.:

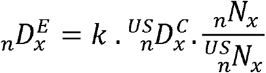

where, separately for males and females, 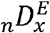 is the global or national number of male/female excess deaths between ages *x* and *x*+*n* in the 12-month period, _*n*_*N*_*x*_ and 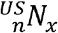 are the number of males/females between ages *x* and *x*+*n* in the world/country and in the U.S.A. on July 1^st^, 2019, 2020 or 2021 (depending on the period) and 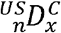 is the male/female number of deaths from COVID-19 between ages *x* and *x*+*n* in the U.S.A. The value of the scalar *k* can be determined so that the sum of 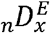 values, across all age groups and both sexes, matches the global or national number of excess deaths. Numbers of males/females by age group in the world and by countries were obtained from the UN projections of national population sizes by sex and age-group for mid-2019, -2020 and -2021 (United Nations 2019). These estimates were regrouped in the same 11 age groups used in the CDC tabulations.

#### Step 3

Sex- and age-specific mortality rates (_*n*_*m*_*x*_) and survival probabilities (_*n*_*p*_*x*_) for the calendar year 2019 and counterfactual rates and probabilities for each of the eight 12-month periods were derived from the UN 2019 estimates and projections. Period values were obtained by linear (_*n*_*m*_*x*_) or exponential (_*n*_*p*_*x*_) interpolation between the 2015-20 and 2020-25 values from the UN.

#### Step 4

excess mortality rates for each country and the world in each of the eight 12-month periods were combined with the counterfactual rates into period life tables by reversing the procedure typically used to “delete” a cause of death from a multiple-decrement life table (Preston, Heuveline, and Guillot 2001; Heuveline and Tzen 2021). This involves calculating new sex- and age-specific survival probabilities for which Chiang (1969) provided an elegant solution:

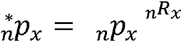

where *p* and _*n*_^*\**^*P*_*x*_ are the survival probabilities for males/females between ages *x* and *x*+*n* in the two life tables and _*n*_*R*_*x*_ is the ratio of projected deaths in the 12-month period with and without excess deaths for males/females in that age interval. This ratio can be obtained as:

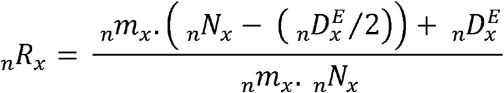

where _*n*_*m*_*x*_ is the male/female age-specific death rate between ages *x* and *x*+*n* in the counterfactual life table for the world/country for this 12-month period (from step 3), and, as above, _*n*_*N*_*x*_ is the mid-2019, -2020 or -2021 global/national population in that sex and age-group, and 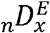 is the global/national number of excess male/female deaths between ages *x* and *x*+*n* in the 12-month period estimated above (step 2). The denominator corresponds to the expected number of deaths without excess mortality, while in the numerator that number is adjusted to take into account the fact that excess deaths modify the exposure to deaths from other causes.

#### Step 5

Life expectancies were estimated from re-estimated probabilities (from step 4) and the counterfactual rates and probabilities (from step 3). To do this in all but the 2019 life table, new values of _*n*_*a*_*x*_, the age-specific number of years lived after age *x* for individuals dying in the age interval *x* to *x*+*n*, also need to be derived, for which the interpolation suggested in Preston et al. (2001, p.84) can be used:

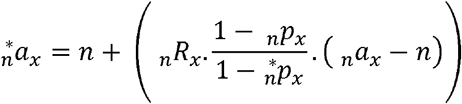

Life expectancies can then be estimated from these new values of _*n*_*a*_*x*_ and the new values of _*n*_*p*_*x*_ (step 5) and standard life table relationships. Values for each period and countries are shown in the Supplementary File “Life Expectancies” (Excel File).

#### Interpreting changes in life expectancy

There are two main interpretations of life expectancy at birth in a period life table. The first one derives from its cohort analog and expresses how long “synthetic” cohort members might be expected to live on average if subjected to unchanged mortality conditions (age-specific mortality rates at every age, or mortality schedule) throughout their lives. The second one is an age-standardized mean age at death in the population, where the actual age distribution is not replaced by an external standard but by its “stationary equivalent,” that is, the age structure of a stationary population with the same mortality schedule. The two interpretations might be linked to the two sides of the life table identity:

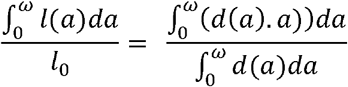

where *l*(*a*) and *d*(*a*) are, respectively, the number of survivors and deaths at exact age *a*. The ratio on the left, total number of years lived, *T*_0_, divided by cohort size, *l*_0_, describes the first interpretation. The ratio on the right describes the second one since

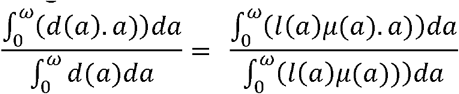

is the mean age at death with the mortality rates by age, *μ*(*a*), and the stationary age distribution *l*(*a*)/*l*_0_, whereas the mean age at death in the population is:

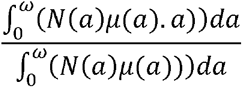

where *N*(a) is the number of individuals of exact age *a* in the population.

The first interpretation is more intuitive and by far the most common. It can be applied to differences in life expectancy between two populations or to the difference made by a mortality change that can be expected to become permanent, such as the elimination of a cause of death. This interpretation becomes problematic when mortality changes temporarily (i.e., during mortality “shocks”) as it would require the assumption that these temporary changes would last throughout the synthetic cohort lifetime.

For these situations, I suggest turning to the second interpretation of life expectancy at birth by interpreting changes in life expectancy as an internally standardized measure of a quantity that might be thought as a “Mean Unfulfilled Lifespan” (Heuveline 2021a). Building on Keyfitz (1977), changes in life expectancy at birth induced by a cause of death, *Δ*^*i*^*e*(0), can be written as:

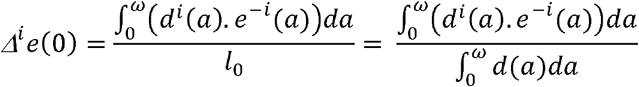

where *d*^*i*^(*a*) is the number of deaths due to cause *i* in the life table and *e*^-*i*^(*a*) is the counterfactual life expectancy at age *a* (in the absence of cause *i*). The difference in life expectancy induced by cause *i* is the stationary equivalent of

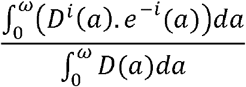

in the population where *D*(*a*) is the number of deaths due to cause *i*. In the numerator we recognize the number of Years of Life Lost due to cause *i*:

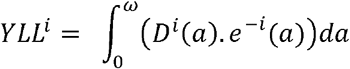

With the total number of deaths during the period, *D*, as a denominator, *YLL*^*i*^/*D* is the average lifespan reduction due to cause *i* among the individuals who died in the reference period. This Mean Unfulfilled Lifespan due to cause *i, MUL*^*i*^ is the product of the average lifespan reduction due to cause *i* for individuals who died of cause *i* times the proportion of all deaths that are due to cause *i* in the reference period:

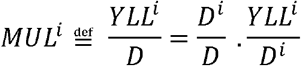

The Mean Unfulfilled Lifespan (MUL) is equally applicable to excess deaths as to deaths from a specific cause and remains interpretable as the number of years by which the average lifespan was cut short by mortality change for individuals who died in the reference period. When mortality changes, the reduction in life expectancy can thus be interpreted as the “stationary-equivalent” of the MUL, regardless of whether the mortality changes can be expected to be permanent or temporary. Again, this interpretation is less intuitive than the most common interpretation of changes in life expectancy, but it is more appropriate when mortality changes are expected to be temporary.

